# Women’s Seclusion during Menstruation and Children’s Health in Nepal

**DOI:** 10.1101/2022.03.18.22272621

**Authors:** Supriya Joshi, Yubraj Acharya

**Author notes:** **Corresponding author:** Department of Agricultural Economics, Sociology, and Education, The Pennsylvania State University, University Park, PA 16802, USA.

## Abstract

**Objectives:** There is limited empirical evidence from low-income countries on the effects of women’s seclusion during menstruation on children’s health. We documented the association between mother’s extreme seclusion during menstruation and children’s health in Nepal.

**Methods:** Using nationally representative data from the 2019 Multiple Indicator Cluster Survey, we examined the relationship between mother’s exposure to extreme forms of seclusion during menstruation and anthropometric measures of nutritional status and health outcomes among children ages 5-59 months (n=6,301). We analyzed the data in a regression framework, controlling for potential confounders, including province fixed effects. We assessed extreme seclusion during menstruation based on women’s exposure to *chhaupadi*, a practice in which women are forced to stay away from home—in separate huts or animal sheds—during menstruation and childbirth.

**Results:** Mothers’ exposure to extreme seclusion during menstruation was associated with 0.18 standard deviation lower height-for-age z-scores (HAZ) (p=0.046) and 0.20 standard deviation lower weight-for-age z-scores (WAZ) (p=0.007) among children. Analysis by the place of seclusion showed that the negative association was stronger when women stayed in animal sheds—0.28 SD for HAZ (p=0.007) and 0.32 SD for WAZ (p<0.001)—than when they stayed in separate huts. Extreme seclusion was associated with higher incidence of acute respiratory symptoms but not with incidence of diarrhea, irrespective of the place of seclusion.

**Conclusions:** Women’s extreme seclusion during menstruation in Nepal has profound implications on the physical health of their children. Additional research is needed to ascertain potential mechanisms.

## Introduction

Violence against women and girls is one of the most widespread and persistent human rights violations in the world today [1], with one in three women subjected to some form of sexual or physical violence [2]. In 2017, the latest year for which data are available, 87,000 women were killed from violence [3]. Existing literature from high-income countries shows that direct violence, such as a mother’s exposure to domestic and intimate-partner violence, adversely affects children’s health [4]—beyond the direct effects on the victims.

In low- and middle-income countries (LMICs), women face many other forms of violence, such as seclusion during menstruation, dowry system, and child marriage. A defining feature of such forms of violence is that they are perpetrated through cultural mechanisms, remain unrecognized, and are unseen because the community normalizes them [5, 6]. The empirical evidence on the effects of such forms of violence is scant. This omission in LMICs is concerning because women are subjected to multiple forms of such violence [7–10]. In these settings, legal and social infrastructure that prevent violence against women, or social protection programs that support victims, are often inadequate or lacking [11–14].

Poor child health and malnutrition is another critical challenge the world faces today. Globally, 144 million children under the age of five suffer from stunting and 47 million children are wasted [15]. UNICEF attributes half of the children’s death under five globally to undernutrition [16]. For surviving children, the adverse consequences of poor nutritional status are likely to persist throughout their lives if malnutrition is not addressed on time [17–20]. These effects manifest in many forms, including reduced cognitive ability [21], increased risk of cardiovascular diseases [22], and disability [23].

Against this background, the objective of the current study was to examine the association between women’s extreme seclusion during menstruation and their children’s nutritional status and health.

## Methods

### Setting

We conducted this study in Nepal, where violence against women is pervasive. To illustrate, 49 gender-based violence related deaths were reported in the first quarter of 2018 (the latest period for which data were available), and 1,874 rape cases were filed with the supreme court between 2016 and 2017 [24]. Women undergo various forms of violence, including child marriage (37% of girls are married by the age of 18 [24]), accusation of practicing witchcraft, dowry-related violence, and trafficking [24].

In the *chhaupadi* practice, which we used as a measure of extreme seclusion, women are secluded and forced to stay separately in makeshift huts or animal sheds during menstruation. The practice has been described in greater detail elsewhere [25–29]. Briefly, the practice is rooted in patriarchy and Hindu mythology that considers women impure, thus untouchable, during menstruation and childbirth. In addition to having to eat and sleep separately, women are not allowed to touch other individuals, eat certain types of food items, visit temples, or worship. The government has attempted to abolish the practice through a series of policies, with limited success. Most recently, in 2017, the government criminalized the practice, specifying fines and jail time for perpetrators through a criminal code. However, the enforcement strategies of this criminal code remain unclear and knowledge about the criminal code is extremely low [29]. The media has frequently reported cases of rape, assault, attack by animals, and suffocation to death of women staying in huts or animal sheds [30–32].

Young children often accompany their mothers during the latter’s seclusion [25, 33], which was the basis for analyzing the relationship between mother’s exposure and child health in the current study. Children in Nepal are among the most vulnerable in the world [34]. Under-five mortality is 25 deaths per 1000 live births [35], and 15% children are born with a low birth weight [36]. The children are severely malnourished, with 36% children below five stunted and 10% wasted [37].

### Data

We used publicly available data from the 2019 Multiple Indicator Cluster Survey (MICS). MICS is a program supported by UNICEF and collects internationally-comparable data on a wide range of indicators on the situation of children and women [38], and is similar to the Demographic and Health Surveys (DHS). Nepal’s 2019 MICS was conducted between May-November 2019. The 2019 survey asked women about their menstruation practices, including whether they had stayed in separate huts or animal sheds during menstruation.

MICS provides separate data files for women and for children ages 0-59 months. For the purpose of this study, we combined the women’s and children’s data files to create a file containing information on 6,301 mother-child pairs. The Stata codes used to combine the data sets, create necessary variables, and conduct the analysis are included in the Online Appendix.

MICS reports that the survey protocol was approved by Nepal’s Central Bureau of Statistics in September 2018 as per the Statistical Act of 1958, and that the statistical act allows CBS to carry out surveys according to the government’s ethics protocol without involving an institutional review board [38]. The current study utilized de-identified, publicly-available data. Therefore, a separate ethical approval was not necessary.

### Measurement of outcome variables

The outcomes for the study were z-scores for height-for-age (HAZ) and weight-for-age (WAZ), incidence of diarrhea, and symptoms of acute respiratory infection (ARI). We chose these outcomes based on data availability and our hypothesized mechanisms through which menstrual seclusion can affect child’s health. The huts where women stay during seclusion are small, often lack a window, and are unsanitary [25, 39]; in most cases, women use the same set of bedding during every menstruation. These conditions can lead to higher incidence of diarrhea and ARI symptoms in young children. When women stay in animal sheds, the children are exposed to animal fecal matter directly. A vast amount of literature has documented the association between exposure to animal feces and children’s health nutritional status [40–42]. Food restrictions imposed on the mother during menstruation can also hinder child’s growth, thus affecting HAZ and WAZ.

MICS used Seca digital scales to measure weight and Shorr measuring boards to record standing/lying height/length of children [43]. HAZ and WAZ for each child are then calculated using the World Health Organization’s reference population [44]. The z-scores reflect how far a child’s height and weight are—in standard deviations—from the median height and weight, respectively, of a healthy reference population. A low HAZ reflects chronic malnutrition—the result of failure to receive adequate nutrition over an extended period. A low WAZ reflects both acute and chronic malnutrition. These z-scores are commonly used to assess children’s nutritional status [45].

For incidences of diarrhea and symptoms of ARI, the questionnaire asked, separately, if the child had diarrhea and breathing problems in the 14 days preceding the survey. According to WHO, diarrhea is the passage of three or more instances of loose stools and a symptom of gastrointestinal infection that usually results from poor hygiene. Diarrhea is often used as a measure of health in studies from LMICs because, if untreated, diarrhea can be life-threatening for young children and is linked with increased morbidity and mortality [46, 47]. For symptoms of ARI, the survey asked the mother if her child had had fast, short, rapid breaths or difficulty breathing in the two weeks preceding the survey [48]. This variable is also a proxy measure of pneumonia, a leading cause of death in children under five [38].

### Measurement of key independent variables

The main predictor variable was a binary measure of whether women had to stay separately during their menstruation. To create this variable, we combined information on whether she stayed in separate huts or in animal shed during menstruation (=1 if she stayed in one of these two places, 0 otherwise). In recent years, an increasing number of women are staying in animal sheds instead of the makeshift huts because of the criminalization of the practice by the government and dismantling of huts by local NGOs [49]. Given this shift, and the potential for differential effects, we also conducted separate analysis by the place of seclusion (huts versus animal shed).

### Measurement of covariates

We included several covariates in our analyses to alleviate concerns about potential confounding. These covariates were selected based on previous evidence on the determinants of children’s nutritional status (specifically, height and weight) and measures of health (diarrhea and symptoms of ARI) [50–52]. Child-specific covariates included the child’s sex, age, and a dietary diversity index. A child’s sex is an important covariate given the prevalent discrimination against girls in the country. The dietary diversity index is also a strong predictor of children’s nutritional status and may be simultaneously correlated with mother’s seclusion during menstruation. MICS asked women what food items they gave their child during the 24 hours preceding the survey. We created the dietary diversity index by grouping the consumed foods into seven categories (staples, legumes, dairy, flesh, vitamin A vegetables, and other vegetables). The index is the total number of these food types.

Mother-specific covariates included the mother’s education, mother’s age at the time of the survey, and mother’s age at first marriage. At the household level, we included household’s access to an improved toilet (binary), access to clean drinking water (binary), and whether household treated the water before drinking (binary). We used the WHO definitions to create these variables [53]. We also controlled for household wealth by including quintiles of the wealth index, available from MICS. Finally, we included a binary measure of whether the household lived in an urban area.

### Statistical analysis

Means and proportions for the outcomes as well as the key household- and child-level characteristics were calculated. To assess the relationship between the outcomes and women seclusion, we estimated the coefficients in a regression of the following form:

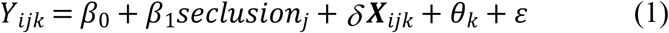

In equation (1), Y_*ijk*_ was the outcome for child *i* of mother *j* living in province *k*. Whether women stayed in a hut or an animal shed during menstruation—the primary independent variable—varied by mother. The coefficient β_1_ reflected the association between this seclusion and the outcome. **X** represented child-, mother-, household- and cluster-level covariates mentioned in the previous subsection. For HAZ and WAZ, we estimated ordinary least square (OLS) regressions.

Given the binary nature of the two other outcomes—incidence of diarrhea and symptoms of ARI—we estimated logistic regressions for these outcomes and reported odds ratios. The logistic regression we estimated took the following form:

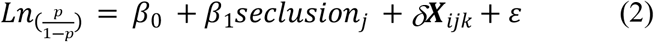

In equation (2), *p* was the probability that a child had incidence of diarrhea (or symptoms of ARI) during the two weeks preceding the survey. The remaining notations carried the same meaning as in equation (1).

Both extreme seclusion during menstruation and the outcomes vary widely across geographic regions in the country. In our analyses, we included province fixed effects to control for potential confounders common to women within a province. Our comparison is thus between children within the same province. A lower-level geographic identifier (such as a district or a municipality) was not available in MICS.

All analysis accounted for the survey design by including the sample weights available from MICS. We carried out all analyses using the Stata software version 16 [54] and reported the statistical significance at the *P*<0.10, *P*<0.05, and *P*<0.01 levels.

## Results

Our analytical sample consisted of 6,301 children ages 0-59 months (Table 1). The average age of the children was 31 months (Table 1, column 1). The average age of the mother was 27 years, with the first marriage occurring at age 19. The majority of women had primary or lower level of education. In the sample, 58% children had access to an improved toilet and 34% had access to clean drinking water. Overall, these characteristics point to the critical influence that having to stay away in huts or animal sheds can have on the health of children. The average WAZ and HAZ scores were -1.24 and -1.39, respectively, suggesting that an average child in the sample had weight and height that were 1.24 and 1.39 standard deviations lower than that of the median child in the WHO’s reference population. One in ten children were reported to have diarrhea during the two weeks preceding the survey, while 7.4% reported having symptoms of ARI.

**Table 1.** Descriptive Statistics for the Analytic Sample.

|  | Total<br>N=6,301 | Did not<br>experience<br>extreme<br>seclusion<br>N=5,923 | Experienced<br>extreme<br>seclusion<br>N=378 | p-value |
| --- | --- | --- | --- | --- |
| **Child characteristics** |  |  |  |  |
| Girl | 47.18% | 47.05% | 49.21% | 0.42 |
| Child's age in months | 30.99 (16.83) | 30.87 (16.90) | 32.9 (15.58) | 0.02 |
| Dietary diversity score | 2.73 (1.91) | 2.71 (1.91) | 3.17 (1.82) | 0.008 |
| **Mother characteristics** |  |  |  |  |
| Mother's age in years | 27.19 (5.78) | 27.19 (5.77) | 27.30 (6.01) | 0.71 |
| Age at first marriage, years | 18.57 (3.81) | 18.61 (3.81) | 17.82 (3.71) | <0.001 |
| Education |  |  |  | <0.001 |
| No education | 23.73% | 22.61% | 41.27% |  |
| Primary | 31.88% | 32.06% | 29.10% |  |
| Secondary | 36.84% | 37.53% | 25.93% |  |
| Post-secondary | 7.55% | 7.80% | 3.70% |  |
| **Household characteristics** |  |  |  |  |
| Access to improved sanitation | 58.48% | 60.44% | 27.78% | <0.001 |
| Access to clean drinking water | 34.15% | 34.81% | 23.81% | <0.001 |
| Water treated before drinking | 76.40% | 75.59% | 89.15% | <0.001 |
| Wealth index quintile |  |  |  | <0.001 |
| Poorest | 28.46% | 25.00% | 82.54% |  |
| Second | 20.55% | 21.34% | 8.20% |  |
| Middle | 19.85% | 20.88% | 3.70% |  |
| Fourth | 18.36% | 19.31% | 3.44% |  |
| Richest | 12.78% | 13.46% | 2.12% |  |
| Urban | 55.58% | 56.49% | 41.27% | <0.001 |
| Ethnicity |  |  |  | <0.001 |
| Brahmin/Chhetri | 32.38% | 29.97% | 70.11% |  |
| Dalit | 17.58% | 17.19% | 23.81% |  |
| Janajati | 30.34% | 32.04% | 3.70% |  |
| Others | 19.70% | 20.80% | 2.38% |  |
| **Outcomes** |  |  |  |  |
| Height for age z-score WHO | -1.39 (1.52) | -1.35 (1.52) | -2.00 (1.37) | <0.001 |
| Weight for age z-score WHO | -1.24 (1.14) | -1.21 (1.14) | -1.79 (1.04) | <0.001 |
| Diarrhea in preceding two weeks | 10.24% | 9.51% | 21.69% | <0.001 |
| Symptoms of acute respiratory infection | 7.36% | 7.02% | 12.70% | <0.001 |

Notes: This table shows the descriptive statistics for the analytic sample. The p-values in the last column are from t-tests (for means) and chi-square tests (for distribution) comparing women who faced extreme seclusion (i.e., stayed in separate huts or animal sheds during menstruation) to those who did not. Source: UNICEF (2019).

Descriptively, children exposed to extreme seclusion differed significantly from those not exposed on most of the characteristics that would matter for children’s health (Table 1, columns 2 and 3). Those exposed were nearly 33 percentage points less likely to have access to an improved toilet and 11 percentage points less likely to have access to clean drinking water. They were disproportionately poor, had lower levels of education, from rural areas, and from the *dalit* groups (who are among the most disadvantaged in the country). In terms of the outcomes, children exposed to extreme seclusion had significantly lower WAZ and HAZ. Specifically, the exposed children had heights that were, on average, 2 standard deviations below the median of the reference population, compared to 1.35 standard deviations below the reference population for children who were not exposed. Likewise, the average weight of the exposed children exposed was 1.79 standard deviations below the median of the reference population, compared to 1.21 standard deviations for children who were not exposed. The exposed children also had higher incidence of both diarrhea (21.7% compared to 9.5%) and ARI symptoms (12.7% compared to 7%) than children who were not exposed.

After adjusting for potential confounders, including the province fixed effects, children exposed to extreme seclusion had 0.18 standard deviations lower HAZ (p=0.046) and 0.20 standard deviations lower WAZ (p=0.007) than that of the children who were not exposed (Table 2, columns 1 and 2). These coefficients suggest that mother’s extreme seclusion during menstruation was associated with 12.9 percent (=100 × 0.18/1.39) and 16.1 (=100 × 0.20/1.24) percent decline in HAZ and WAZ of their children, respectively, at the respective means.

**Table 2.**
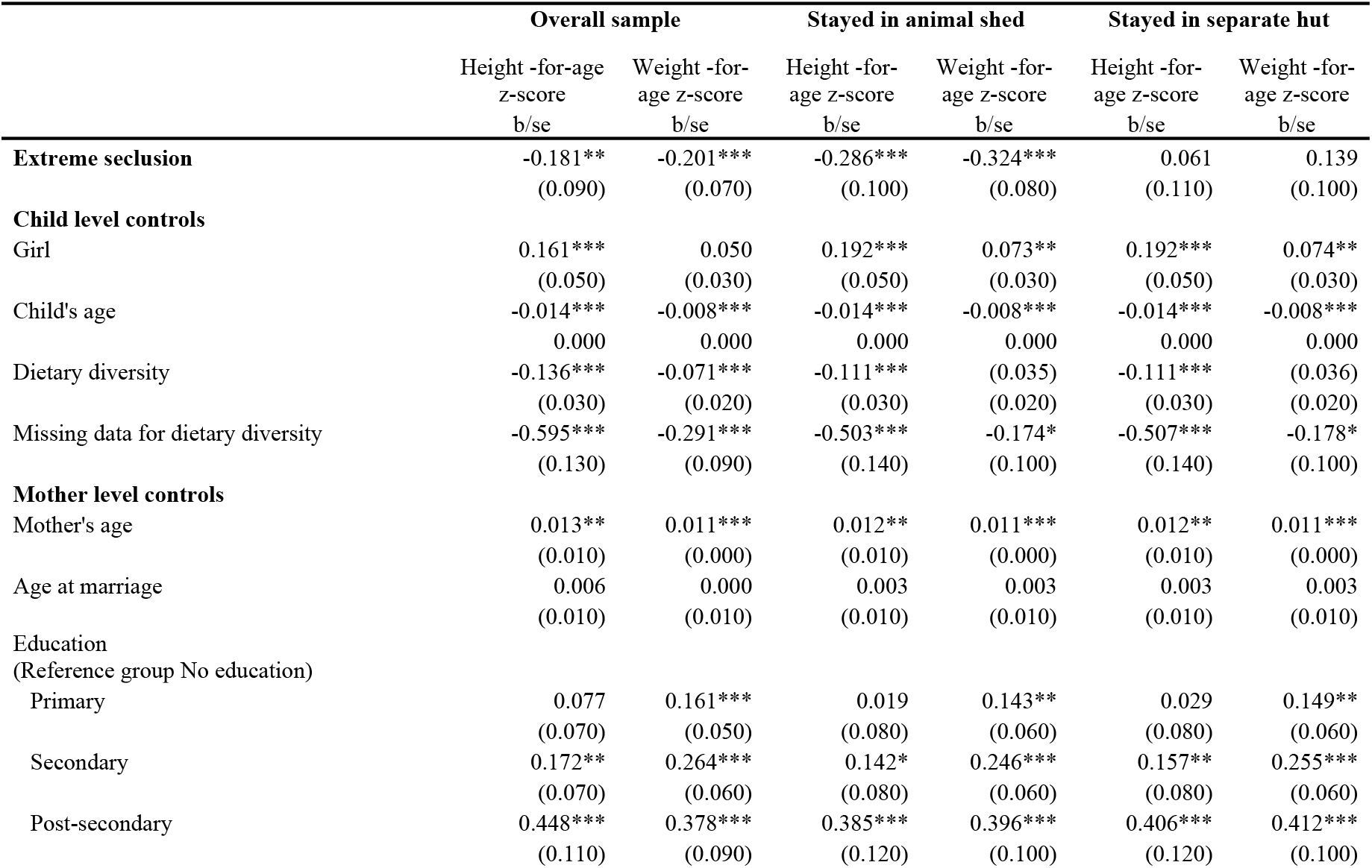
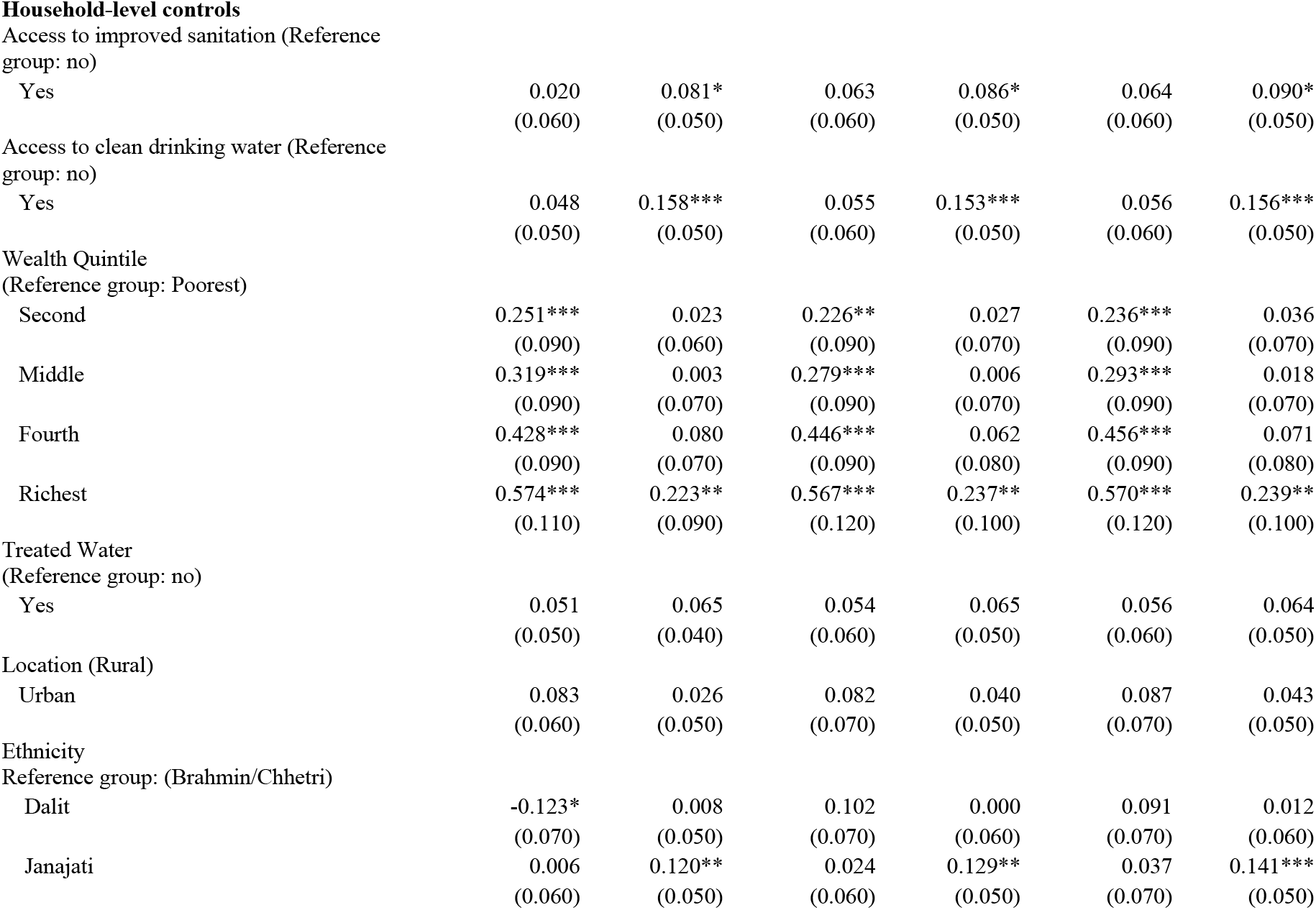
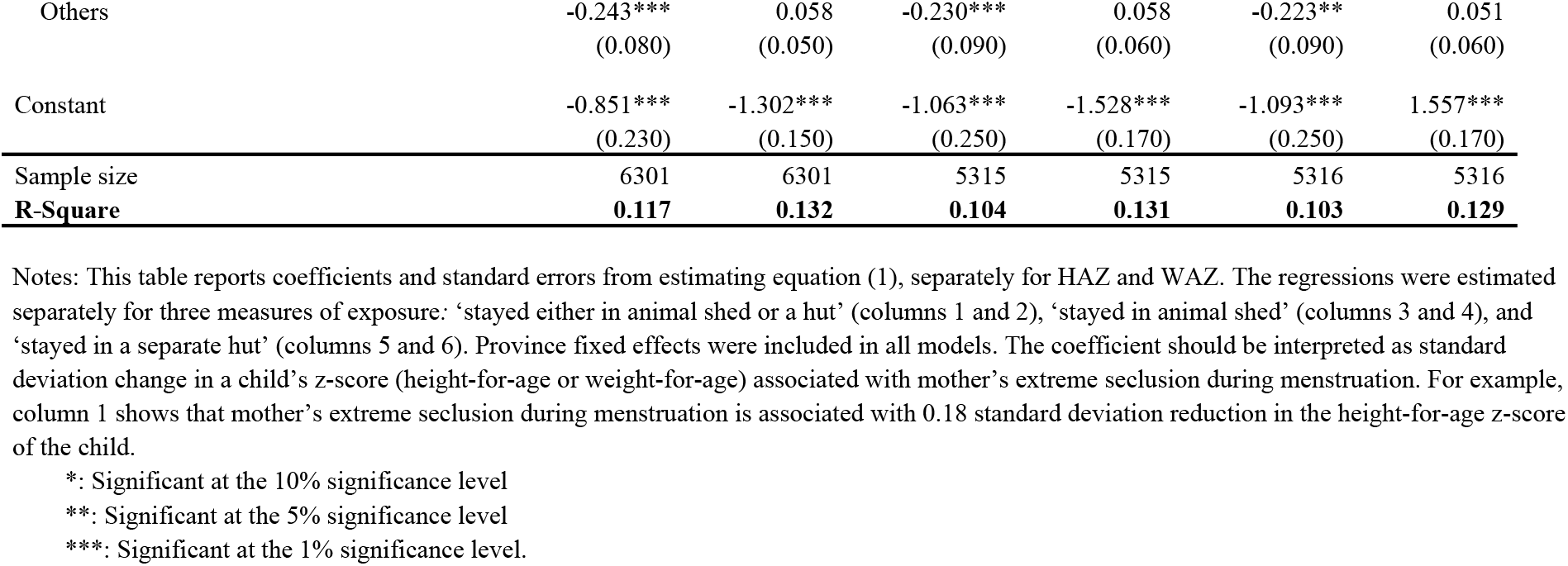
Coefficients from Linear Regressions of Height-for-Age and Weight-for-Age Z-scores of Children 5-59 Months on Whether the Mother Stayed Separately in Animal Shed or Hut during Menstruation.

|  | Overall sample |  | Stayed in animal shed |  | Stayed in separate hut |  |
| --- | --- | --- | --- | --- | --- | --- |
|  | Height -for-age<br>z-score<br>b/se | Weight -for-age z-score<br>b/se | Height -for-age z-score<br>b/se | Weight -for-age z-score<br>b/se | Height -for-age z-score<br>b/se | Weight -for-age z-score<br>b/se |
| **Extreme seclusion** | -0.181**<br>(0.090) | -0.201***<br>(0.070) | -0.286***<br>(0.100) | -0.324***<br>(0.080) | 0.061<br>(0.110) | 0.139<br>(0.100) |
| **Child level controls** |  |  |  |  |  |  |
| Girl | 0.161***<br>(0.050) | 0.050<br>(0.030) | 0.192***<br>(0.050) | 0.073**<br>(0.030) | 0.192***<br>(0.050) | 0.074**<br>(0.030) |
| Child's age | -0.014***<br>0.000 | -0.008***<br>0.000 | -0.014***<br>0.000 | -0.008***<br>0.000 | -0.014***<br>0.000 | -0.008***<br>0.000 |
| Dietary diversity | -0.136***<br>(0.030) | -0.071***<br>(0.020) | -0.111***<br>(0.030) | (0.035)<br>(0.020) | -0.111***<br>(0.030) | (0.036)<br>(0.020) |
| Missing data for dietary diversity | -0.595***<br>(0.130) | -0.291***<br>(0.090) | -0.503***<br>(0.140) | -0.174*<br>(0.100) | -0.507***<br>(0.140) | -0.178*<br>(0.100) |
| **Mother level controls** |  |  |  |  |  |  |
| Mother's age | 0.013**<br>(0.010) | 0.011***<br>(0.000) | 0.012**<br>(0.010) | 0.011***<br>(0.000) | 0.012**<br>(0.010) | 0.011***<br>(0.000) |
| Age at marriage | 0.006<br>(0.010) | 0.000<br>(0.010) | 0.003<br>(0.010) | 0.003<br>(0.010) | 0.003<br>(0.010) | 0.003<br>(0.010) |
| Education<br>(Reference group No education) |  |  |  |  |  |  |
| Primary | 0.077<br>(0.070) | 0.161***<br>(0.050) | 0.019<br>(0.080) | 0.143**<br>(0.060) | 0.029<br>(0.080) | 0.149**<br>(0.060) |
| Secondary | 0.172**<br>(0.070) | 0.264***<br>(0.060) | 0.142*<br>(0.080) | 0.246***<br>(0.060) | 0.157**<br>(0.080) | 0.255***<br>(0.060) |
| Post-secondary | 0.448***<br>(0.110) | 0.378***<br>(0.090) | 0.385***<br>(0.120) | 0.396***<br>(0.100) | 0.406***<br>(0.120) | 0.412***<br>(0.100) |
| Yes | 0.020<br>(0.060) | 0.081*<br>(0.050) | 0.063<br>(0.060) | 0.086*<br>(0.050) | 0.064<br>(0.060) | 0.090*<br>(0.050) |
| Access to clean drinking water (Reference group: no) |  |  |  |  |  |  |
| Yes | 0.048<br>(0.050) | 0.158***<br>(0.050) | 0.055<br>(0.060) | 0.153***<br>(0.050) | 0.056<br>(0.060) | 0.156***<br>(0.050) |
| Second | 0.251***<br>(0.090) | 0.023<br>(0.060) | 0.226**<br>(0.090) | 0.027<br>(0.070) | 0.236***<br>(0.090) | 0.036<br>(0.070) |
| Middle | 0.319***<br>(0.090) | 0.003<br>(0.070) | 0.279***<br>(0.090) | 0.006<br>(0.070) | 0.293***<br>(0.090) | 0.018<br>(0.070) |
| Fourth | 0.428***<br>(0.090) | 0.080<br>(0.070) | 0.446***<br>(0.090) | 0.062<br>(0.080) | 0.456***<br>(0.090) | 0.071<br>(0.080) |
| Richest | 0.574***<br>(0.110) | 0.223**<br>(0.090) | 0.567***<br>(0.120) | 0.237**<br>(0.100) | 0.570***<br>(0.120) | 0.239**<br>(0.100) |
| Yes | 0.051<br>(0.050) | 0.065<br>(0.040) | 0.054<br>(0.060) | 0.065<br>(0.050) | 0.056<br>(0.060) | 0.064<br>(0.050) |
| Location (Rural) |  |  |  |  |  |  |
| Urban | 0.083<br>(0.060) | 0.026<br>(0.050) | 0.082<br>(0.070) | 0.040<br>(0.050) | 0.087<br>(0.070) | 0.043<br>(0.050) |
| Dalit | -0.123*<br>(0.070) | 0.008<br>(0.050) | 0.102<br>(0.070) | 0.000<br>(0.060) | 0.091<br>(0.070) | 0.012<br>(0.060) |
| Janajati | 0.006<br>(0.060) | 0.120**<br>(0.050) | 0.024<br>(0.060) | 0.129**<br>(0.050) | 0.037<br>(0.070) | 0.141***<br>(0.050) |
| Others | -0.243***<br>(0.080) | 0.058<br>(0.050) | -0.230***<br>(0.090) | 0.058<br>(0.060) | -0.223**<br>(0.090) | 0.051<br>(0.060) |
| Constant | -0.851***<br>(0.230) | -1.302***<br>(0.150) | -1.063***<br>(0.250) | -1.528***<br>(0.170) | -1.093***<br>(0.250) | 1.557***<br>(0.170) |
| Sample size | 6301 | 6301 | 5315 | 5315 | 5316 | 5316 |
| **R-Square** | **0.117** | **0.132** | **0.104** | **0.131** | **0.103** | **0.129** |

Notes: This table reports coefficients and standard errors from estimating equation (1), separately for HAZ and WAZ. The regressions were estimated separately for three measures of exposure: ‘stayed either in animal shed or a hut’ (columns 1 and 2), ‘stayed in animal shed’ (columns 3 and 4), and ‘stayed in a separate hut’ (columns 5 and 6). Province fixed effects were included in all models. The coefficient should be interpreted as standard deviation change in a child’s z-score (height-for-age or weight-for-age) associated with mother’s extreme seclusion during menstruation. For example, column 1 shows that mother’s extreme seclusion during menstruation is associated with 0.18 standard deviation reduction in the height-for-age z-score of the child.
\*: Significant at the 10% significance level
\*\*: Significant at the 5% significance level
\*\*\*: Significant at the 1% significance level.

The overall association between extreme seclusion during menstruation and the outcomes masked significant heterogeneity in associations based on the nature of seclusion (i.e., staying in huts verse animal sheds). Specifically, children of women who were forced to stay in animal sheds had HAZ and WAZ that were 0.29 and 0.32 standard deviations lower (p=0.007 and p<0.001, respectively) than that of children whose mothers did not face extreme seclusion (columns 3 and 4). In contrast, staying in a separate hut was not associated with either of these outcomes (columns 5 and 6).

Mother’s extreme seclusion during menstruation was significantly associated with higher odds of ARI among children, irrespective of the nature of seclusion (Table 3; aOR=2.19, p=0.01 for overall; aOR=2.48, p=0.003 when women stayed in animal shed; and aOR=2.46, p=0.006 when women stayed in separate huts). Substantively, children whose mothers faced extreme seclusion had more than two-fold increase in the chances of having symptoms of ARI.

**Table 3.** Odds Ratios from Logistic Regressions of Incidence of Diarrhea and Symptoms of Acute Respiratory Infection (ARI) among Children 5-59 Months on Whether the Mother Stayed Separately in Animal Shed or Hut during Menstruation.

|  | **Overall sample** |  | **Stayed in animal shed** |  | **Stayed in separate hut** |  |
| --- | --- | --- | --- | --- | --- | --- |
|  | Diarrhea<br>b/se | Symptoms of ARI<br>b/se | Diarrhea<br>b/se | Symptoms of ARI<br>b/se | Diarrhea<br>b/se | Symptoms of ARI<br>b/se |
| **Extreme seclusion** | 1.361<br>(0.310) | 2.118**<br>(0.620) | 1.517<br>(0.410) | 2.483***<br>(0.750) | 1.415<br>(0.400) | 2.458***<br>(0.810) |
| **Child level controls** |  |  |  |  |  |  |
| Girl | 0.771**<br>(0.090) | 0.809<br>(0.110) | 0.776**<br>(0.100) | 0.797*<br>(0.110) | 0.777**<br>(0.100) | 0.802<br>(0.110) |
| Child's age | 0.990*** | 0.985*** | 0.988*** | 0.983*** | 0.988*** | 0.983*** |
| **Mother level controls** |  |  |  |  |  |  |
| Mother's age | 0.980*<br>(0.010) | 0.985<br>(0.010) | 0.983<br>(0.010) | 0.986<br>(0.010) | 0.982<br>(0.010) | 0.986<br>(0.010) |
| Age at marriage | 1.020<br>(0.020) | 1.011<br>(0.020) | 1.023<br>(0.020) | 1.017<br>(0.020) | 1.022<br>(0.020) | 1.018<br>(0.020) |
| Education (Reference group: no education) |  |  |  |  |  |  |
| Primary | 0.999<br>(0.160) | 1.127<br>(0.190) | 1.042<br>(0.190) | 0.980<br>(0.180) | 1.039<br>(0.190) | 0.977<br>(0.180) |
| Secondary | 0.625***<br>(0.110) | 1.074<br>(0.230) | 0.662**<br>(0.130) | 1.009<br>(0.230) | 0.657**<br>(0.130) | 0.997<br>(0.230) |
| Post-secondary | 0.794<br>(0.210) | 1.015<br>(0.380) | 0.700<br>(0.220) | 0.999<br>(0.410) | 0.718<br>(0.220) | 1.016<br>(0.410) |
| Access to improved sanitation (Reference group: no) |  |  |  |  |  |  |
| Yes | 0.875<br>(0.110) | 0.705**<br>(0.120) | 0.892<br>(0.130) | 0.722*<br>(0.130) | 0.887<br>(0.130) | 0.719*<br>(0.130) |
| Access to clean drinking water (Reference group: no) |  |  |  |  |  |  |
| Yes | 0.894<br>(0.110) | 1.273<br>(0.200) | 0.822<br>(0.120) | 1.182<br>(0.180) | 0.822<br>(0.120) | 1.194<br>(0.180) |
| **Household-level controls** |  |  |  |  |  |  |
| Wealth Quintile (Reference group: poorest) |  |  |  |  |  |  |
| Second | 0.878<br>(0.150) | 0.801<br>(0.160) | 0.759<br>(0.140) | 0.757<br>(0.160) | 0.753<br>(0.140) | 0.761<br>(0.160) |
| Middle | 0.617**<br>(0.120) | 0.930<br>(0.220) | 0.525***<br>(0.110) | 0.821<br>(0.220) | 0.527***<br>(0.110) | 0.829<br>(0.220) |
| Fourth | 0.936<br>(0.210) | 0.780<br>(0.200) | 0.779<br>(0.200) | 0.676<br>(0.180) | 0.789<br>(0.200) | 0.694<br>(0.190) |
| Richest | 0.682<br>(0.190) | 0.994<br>(0.370) | 0.686<br>(0.210) | 1.008<br>(0.400) | 0.685<br>(0.210) | 1.015<br>(0.410) |
| Treated Water (Reference group: no) |  |  |  |  |  |  |
| Yes | 0.718**<br>(0.100) | 1.037<br>(0.170) | 0.734*<br>(0.120) | 1.068<br>(0.180) | 0.726**<br>(0.120) | 1.055<br>(0.170) |
| Location (Reference group: Rural) |  |  |  |  |  |  |
| Urban | 0.786*<br>(0.100) | 0.887<br>(0.160) | 0.759*<br>(0.110) | 0.866<br>(0.150) | 0.763*<br>(0.110) | 0.867<br>(0.150) |
| Ethnicity ((Reference group: Brahmin/Chhetri) |  |  |  |  |  |  |
| Dalit | 1.148<br>(0.190) | 1.115<br>(0.250) | 1.278<br>(0.230) | 1.262<br>(0.320) | 1.257<br>(0.220) | 1.234<br>(0.310) |
| Janajati | 1.032<br>(0.180) | 0.760<br>(0.170) | 1.073<br>(0.210) | 0.817<br>(0.190) | 1.063<br>(0.200) | 0.812<br>(0.190) |
| Others | 1.313<br>(0.250) | 0.642<br>(0.190) | 1.265<br>(0.260) | 0.737<br>(0.250) | 1.244<br>(0.250) | 0.729<br>(0.250) |
| Sample size | 6301 | 6301 | 5315 | 5315 | 5316 | 5316 |

Notes: This table reports odds ratios and standard errors from estimating equation (1), separately for HAZ and WAZ. The regressions were estimated separately for three measures of exposure: ‘stayed either in animal shed or a hut’ (columns 1 and 2), ‘stayed in animal shed’ (columns 3 and 4), and ‘stayed in a separate hut’ (columns 5 and 6). Province fixed effects were included in all models.
\*: Significant at the 10% significance level
\*\*: Significant at the 5% significance level
\*\*\*: Significant at the 1% significance level.

Contrary to expectation, extreme seclusion was not associated with incidence of diarrhea and this lack of association did not vary by the nature of seclusion (aOR=1.36, p=0.18 for overall; aOR=1.52, p=0.12 when women stayed in animal shed; and aOR=1.41, p=0.21 when women stayed in separate huts). We return to this surprising finding in the Discussion.

## Discussion and Conclusion

In this study, we examined the association between extreme seclusion during menstruation and their children’s health. We found, first, that mother’s extreme seclusion was strongly associated with worse nutritional outcomes for children. The limited literature on the effect of mother’s exposure to similar forms of violence on children’s health outcomes makes it difficult to situate our findings in the extant literature. In fact, our previous review suggests that the health effects of the Chhaupadi practice in Nepal has not been examined before [55]. However, our findings agree with the literature on the effects of *direct* forms of violence, such as sexual violence, on children’s health [56–60].

In our study, the consequences for children were more adverse when the mother had to stay in animal sheds than in separate huts. One possible explanation for this difference is that the huts are cleaner than animal sheds, thus exposure to pathogens is lower in the huts. In fact, we found no association between whether the mother stayed in a hut and her children’s HAZ and WAZ. This finding has important policy implications because many women have reportedly shifted to animal sheds from huts after the government criminalized the practice of staying in separate huts in 2017. It appears that, with this shift, children’s health can get worse than when women stayed in huts.

Second, children whose mothers who faced extreme seclusion were significantly more likely to have symptoms of ARI than those whose mothers were not exposed. Previous qualitative studies have documented that women frequently complain about not having enough bedding materials, warm clothes, and blankets during seclusion. During winter months, children are exposed to extreme cold conditions for 4-6 nights. Those studies have also reported that small children had to be bathed in an open area for “purification” before they could get into their house, further exposing them to the cold. Previous studies have found such exposures to the cold be associated with respiratory infections and pneumonia [61, 62].

The lack of an association between extreme seclusion and incidence of diarrhea, particularly when the mother was forced to stay in an animal shed, is striking, as previous studies have established that exposure to animal and animal feces leads to many diseases including diarrhea [63–65]. One possible explanation for this surprising finding is methodological. Specifically, the lack of statistical significance in our study despite large, substantive association raises concerns about the potential lack of statistical power. Descriptively (Table 1), the difference in the incidence of diarrhea between children whose mothers faced extreme seclusion and those who did not was higher than the difference in the incidence of ARI. From the regression, the odds ratios on extreme seclusion are smaller for diarrhea than for ARI (Table 3). However, they are still large substantively—staying in shed is associated with nearly a 50% increase in the odds of diarrhea incidence (Table 3, column 3).

The reader should understand our findings in light of a number of additional limitations. First, given the cross-sectional nature of the data, we could not establish causality. Although we controlled for several potential confounders, some of the observed association between extreme seclusion and our outcomes could be due to residual confounding. Second, our exposure variable was based on women’s self-report and susceptible to under-reporting. Our estimates are biased downward if women who faced extreme seclusion but reported not facing it have children with poorer nutritional status and health. Conversely, our estimates are biased upward if such women have children with better nutritional status and health. It is difficult to ascertain the overall direction of the bias. Finally, the outcomes we evaluated are only a few among the many that extreme seclusion during menstruation can affect; our outcomes were limited by data availability.

In addition to addressing these limitations, future research can delve deeper into the mechanisms through which seclusion during menstruation can affect the health of mothers and children. These mechanisms may include changes in pre- and post-natal behavior, fetal growth, and care practices, among others [4]. Given data limitations, we were unable to assess these mechanisms. Additional data—including on *how* the seclusion takes place (e.g., duration of stay and specifics of sanitary practices) and effect on mothers themselves (e.g., on mental health)— will be required to understand the full extent of the effects on children’s health and to design targeted interventions to reduce these effects. Longer-term, of course, the policy goal has to be to eliminate seclusion during menstruation altogether even if there are no health consequences—as addressing the violation of women’s human right is an important goal in itself.

## Data Availability

This study uses publicly-available, de-identified data from the Multiple Indicator Cluster Surveys (MICS). These data can be downloaded from https://mics.unicef.org free of charge (requires registration).

## Acknowledgement

We would like to thank Heather Randall for comments on an earlier draft of this paper and UNICEF for making the 2019 MICS data available. All errors are our own.

## Data availability

This study uses publicly-available, de-identified data from the Multiple Indicator Cluster Surveys (MICS). These data can be downloaded from https://mics.unicef.org free of charge (requires registration). The Stata codes used to compile the analytic sample and conduct the analysis are included as Online Supplement to this manuscript.

## Patient consent statement

Ethics approval was not required for this paper because it uses publicly-available, de-identified data from the Multiple Indicator Cluster Surveys (MICS; https://mics.unicef.org/). MICS reports that the survey protocol was approved by Nepal’s Central Bureau of Statistics (CBS) in September 2018 as per the Statistical Act of 1958, and that the Statistical Act allows CBS to carry out surveys according to the government’s ethics protocol without involving an institutional review board.

